# HIGHER LEVELS OF CEREBROSPINAL FLUID NITRITE AND NITRATE IN LEVODOPA-INDUCED DYSKINESIAS IN PARKINSON’S DISEASE

**DOI:** 10.1101/2020.06.04.20122283

**Authors:** Bruno L. Santos-Lobato, Mariza Bortolanza, Marcelo E. Batalhão, Ângela V. Pimentel, Evelin Capellari-Carnio, Elaine A. Del-Bel, Vitor Tumas

## Abstract

**Introduction:** Levodopa-induced dyskinesias (LID) in Parkinson’s disease (PD) are frequent complications, and nitric oxide has a role on its pathophysiology. The present work aims to investigate CSF levels of nitric oxide metabolites nitrite and nitrate (NOx) in patients with PD and LID.

**Methods:** We measured CSF NOx levels in patients with PD with and without LID, and in healthy controls. The levels of CSF NOx levels were measured by ozone-based chemiluminescence.

**Results:** 67 participants were enrolled. CSF NOx levels were higher in patients with PD with LID than in healthy controls (Kruskal-Wallis statistics = 7.24, p = 0.02). CSF NOx levels did not correlate with other clinical variables.

**Conclusion:** We reported higher levels of nitric oxide in the CSF of patients with PD and LID.

**Highlights:**

– Nitric oxide has a role on levodopa-induced dyskinesias in Parkinson’s disease
– We measured CSF nitrite and nitrate in Parkinson’s disease patients with dyskinesias
– CSF nitrite and nitrate were higher in Parkinson’s disease patients with dyskinesias

## 1. Introduction

Parkinson’s disease (PD) is the second most common neurodegenerative disease, and most patients develop motor complications associated with the chronic use of levodopa, including levodopa-induced dyskinesias (LID). LID affect 33 to 51.2% of patients with PD after five years of levodopa therapy [1].

A growing number of evidences have shown the role of nitric oxide (NO) on motor control through its interactions with the nigrostriatal pathways, and on the pathophysiology of LID [2]. Pharmacological inhibition of NO generation by compounds as 7-nitroindazole, L-NOARG and L-NAME attenuated LID in hemi-Parkinsonian rats submitted to levodopa therapy [3].

The basal ganglia nitrergic system has been described in healthy human brains [4] and in brains of patients with PD [5]. Nitrergic activity in PD has also been explored in cerebrospinal fluid (CSF) of patients with the measurement of their metabolites, nitrite and nitrate [6, 7, 8, 9, 10, 11]. No previous study assessed the association of NO with LID in patients with PD.

To confirm the role of NO on the pathophysiology of LID in PD, we conducted a study to investigate the association of CSF levels of NO metabolites nitrite and nitrate (NOx) with LID in patients with PD.

## 2. Materials and Methods

### 2.1. Study design and participants

We conducted an observational cross-sectional study to analyze NOx levels in CSF of patients with PD with and without LID and in healthy controls. Participants were recruited in Movement Disorders Unit of Ribeirão Preto Medical School, Brazil, between October 2015 and May 2017. All patients with PD met the UK Parkinson’s Disease Society Brain Bank clinical diagnostic criteria for PD. Healthy controls were enrolled from a volunteers cohort. Patients with PD and healthy controls were included using a 1:1 ratio matching for sex and age within 4 years.

We excluded participants if they had: (1) acute or chronic infections, severe systemic conditions, autoimmune disorders or other neurological diseases; (2) kidney or chronic liver diseases or regular alcohol intake (over 80 g/day for 6 months); (3) high nitrite/nitrate dietary intake (as strict vegetarian or vegan diet); (4) intake of drugs containing nitrite/nitrate. Additionally, Patients with PD were excluded if they had: (1) PD dementia, or (2) mild psychosis defined as a score > 1 on item 1.2 of the International Parkinson and Movement Disorders Society – Unified Parkinson’s Disease Rating Scale (MDS-UPDRS) Part I (hallucinations and psychosis).

For analysis, participants were divided into three groups: healthy controls (HC), patients with PD without LID (PD-ND) and patients with PD with LID (PD-D). The study was approved by the Ribeirão Preto Medical School Ethics Committee (Number 3.036.243), and all participants provided written informed consent.

### 2.2. Evaluations

All patients with PD were examined by the same movement disorder specialist (B.L.S.L.) using a standardized assessment including the MDS-UPDRS [12] and the Unified Dyskinesia Rating Scale (UDysRS) [13] in the ON-stage for patients with LID. We defined the presence of LID by a score ≥ 1 on item 4.1 of the MDS-UPDRS Part IV (time spent with dyskinesias) and confirmed if patient presented abnormal movements in ON-stage.

### 2.3. Collection, processing and storage of biologic samples

We collected CSF between 08:00 and 10:00 AM, without fasting. Patients were instructed to take their morning dose of levodopa. CSF samples were placed on ice immediately after collection. For CSF collection, we performed lumbar puncture, in lateral recumbent position, using a traumatic Quincke (0.7 mm X 63 mm) needle at the L3/L4 or L4/L5 level. We separated 1–2 mL of CSF for routine analysis (cell counts, glucose, proteins). All remaining CSF (10–12 mL) was collected in polyprolylene tubes and gently mixed to avoid gradient effects, centrifuged at 4°C at 4000 g for 10 minutes to remove cells, aliquoted into 1-mL cryotubes, coded, and after stored at –80°C until use without preservatives added. CSF samples contaminated with blood (CSF red cells > 500 per mm^3^) were excluded.

### 2.4. Measurement of CSF nitrite and nitrate levels

First, 50 µL aliquots of CSF were deproteinated by precipitation before measurement using 100 µL 100% ethanol at 4°C, followed by agitation and resting for 30 minutes in freezer –20°C, and centrifuged at 25°C at 4000 g for 10 minutes. NOx levels were measured by ozone-based chemiluminescence using a Sievers® Nitric Oxide Analyzer 280 (GE Analytical Instruments, Boulder, USA). For measurement, 5 µL of prepared samples were injected into the purge vessel with a reducing agent (0.8% vanadium (III) chloride in 1N hydrochloric acid at 95°C), converting nitrate to NO. NO is carried through NO analyzer, which generates ozone to react with NO, forming nitrogen dioxide and chemiluminescence. Emitted photon is detected by the photomultiplier tube, converted to electric signal, which is amplified and recorded by a specific software [14]. NOx levels were reported in µM.

### 2.5. Statistical analysis

To compare two independent groups, we used the Mann-Whitney test; to compare three or more groups, we performed Kruskal-Wallis test for continuous variables (followed by Dunn’s test for multiple comparisons), and the chi-square test for categorical variables. To compare two continuous variables, we performed the Spearman’s Rho correlation test. All analyses were performed using SPSS for Windows version 23.0 (SPSS Inc., Chicago, USA) and figures were made using GraphPrism for Windows version 5.0 (GraphPad Software Inc., La Jolla, USA).

## 3. Results

### 3.1. Clinical and epidemiological data

We recruited a total of 76 participants for this study. Of these, 67 participants fulfilled inclusion and exclusion criteria. We did not perform CSF analysis in 4 patients with PD (with LID – 1 patient, without LID – 3 patients) due to technical problems with lumbar puncture. Data are summarized in Table 1.

**Table 1.**
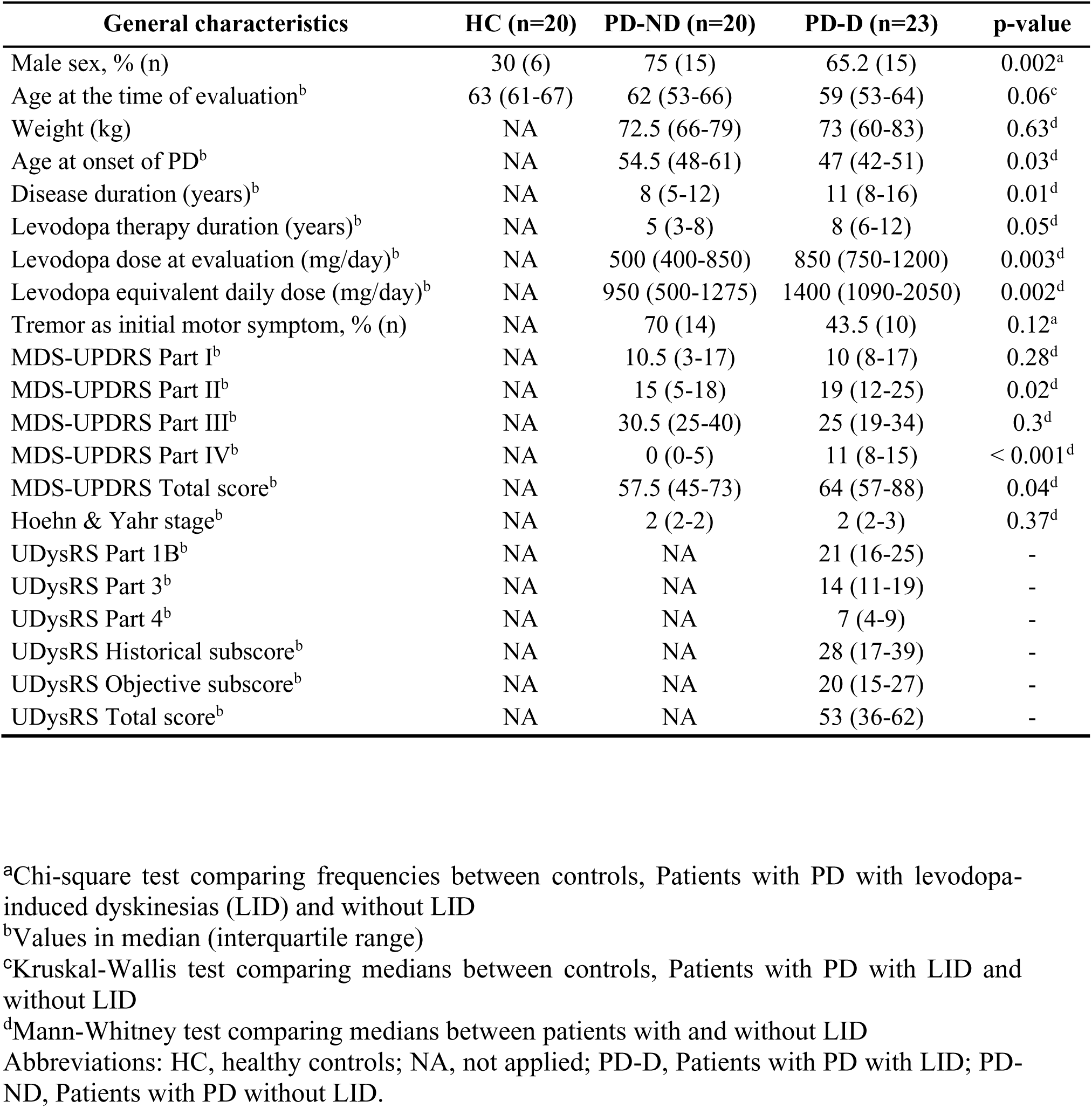
Clinical and epidemiological data of patients with Parkinson’s disease (PD) and healthy controls.

| General characteristics | HC (n=20) | PD-ND (n=20) | PD-D (n=23) | p-value |
| --- | --- | --- | --- | --- |
| Male sex, % (n) | 30 (6) | 75 (15) | 65.2 (15) | 0.002<sup>a</sup> |
| Age at the time of evaluation<sup>b</sup> | 63 (61-67) | 62 (53-66) | 59 (53-64) | 0.06<sup>c</sup> |
| Weight (kg) | NA | 72.5 (66-79) | 73 (60-83) | 0.63<sup>d</sup> |
| Age at onset of PD<sup>b</sup> | NA | 54.5 (48-61) | 47 (42-51) | 0.03<sup>d</sup> |
| Disease duration (years)<sup>b</sup> | NA | 8 (5-12) | 11 (8-16) | 0.01<sup>d</sup> |
| Levodopa therapy duration (years)<sup>b</sup> | NA | 5 (3-8) | 8 (6-12) | 0.05<sup>d</sup> |
| Levodopa dose at evaluation (mg/day)<sup>b</sup> | NA | 500 (400-850) | 850 (750-1200) | 0.003<sup>d</sup> |
| Levodopa equivalent daily dose (mg/day)<sup>b</sup> | NA | 950 (500-1275) | 1400 (1090-2050) | 0.002<sup>d</sup> |
| Tremor as initial motor symptom, % (n) | NA | 70 (14) | 43.5 (10) | 0.12<sup>a</sup> |
| MDS-UPDRS Part I<sup>b</sup> | NA | 10.5 (3-17) | 10 (8-17) | 0.28<sup>d</sup> |
| MDS-UPDRS Part II<sup>b</sup> | NA | 15 (5-18) | 19 (12-25) | 0.02<sup>d</sup> |
| MDS-UPDRS Part III<sup>b</sup> | NA | 30.5 (25-40) | 25 (19-34) | 0.3<sup>d</sup> |
| MDS-UPDRS Part IV<sup>b</sup> | NA | 0 (0-5) | 11 (8-15) | < 0.001<sup>d</sup> |
| MDS-UPDRS Total score<sup>b</sup> | NA | 57.5 (45-73) | 64 (57-88) | 0.04<sup>d</sup> |
| Hoehn & Yahr stage<sup>b</sup> | NA | 2 (2-2) | 2 (2-3) | 0.37<sup>d</sup> |
| UDysRS Part 1B<sup>b</sup> | NA | NA | 21 (16-25) | - |
| UDysRS Part 3<sup>b</sup> | NA | NA | 14 (11-19) | - |
| UDysRS Part 4<sup>b</sup> | NA | NA | 7 (4-9) | - |
| UDysRS Historical subscore<sup>b</sup> | NA | NA | 28 (17-39) | - |
| UDysRS Objective subscore<sup>b</sup> | NA | NA | 20 (15-27) | - |
| UDysRS Total score<sup>b</sup> | NA | NA | 53 (36-62) | - |

<sup>a</sup>Chi-square test comparing frequencies between controls, Patients with PD with levodopa-induced dyskinesias (LID) and without LID
<sup>b</sup>Values in median (interquartile range)
<sup>c</sup>Kruskal-Wallis test comparing medians between controls, Patients with PD with LID and without LID
<sup>d</sup>Mann-Whitney test comparing medians between patients with and without LID
Abbreviations: HC, healthy controls; NA, not applied; PD-D, Patients with PD with LID; PD-ND, Patients with PD without LID.

There was no difference in age at the time of evaluation between groups, but sex frequency was different between patients with PD and healthy controls. Between patients with PD, the PD-D group had earlier onset of disease, longer disease and levodopa therapy duration, and higher doses of antiparkinsonian drugs; however, motor symptoms and staging were similar between groups.

### 3.2. CSF nitrite and nitrate levels in different groups

CSF NOx levels are shown in Fig. 1A (HC group: median 12.4 μM, interquartile range 10.1–15; PD-ND group: median 12.7 μM, interquartile range 10.7–16.9; PD-D group: 17.5 μM, interquartile range 13–26.1). CSF NOx levels in PD-D group were higher than in healthy controls (Kruskal-Wallis statistics = 7.24, p = 0.02 – HC group versus PD-D group, difference in rank sum = 14.17, p < 0.05). Even with a more severe disease in PD-D group, covariance analysis did not show a significant effect of age at onset, disease duration, levodopa therapy duration and antiparkinsonian drugs doses on CSF NOx levels between with and without LID PD groups (adjusted p-value = 0.12).

**Figure 1.**
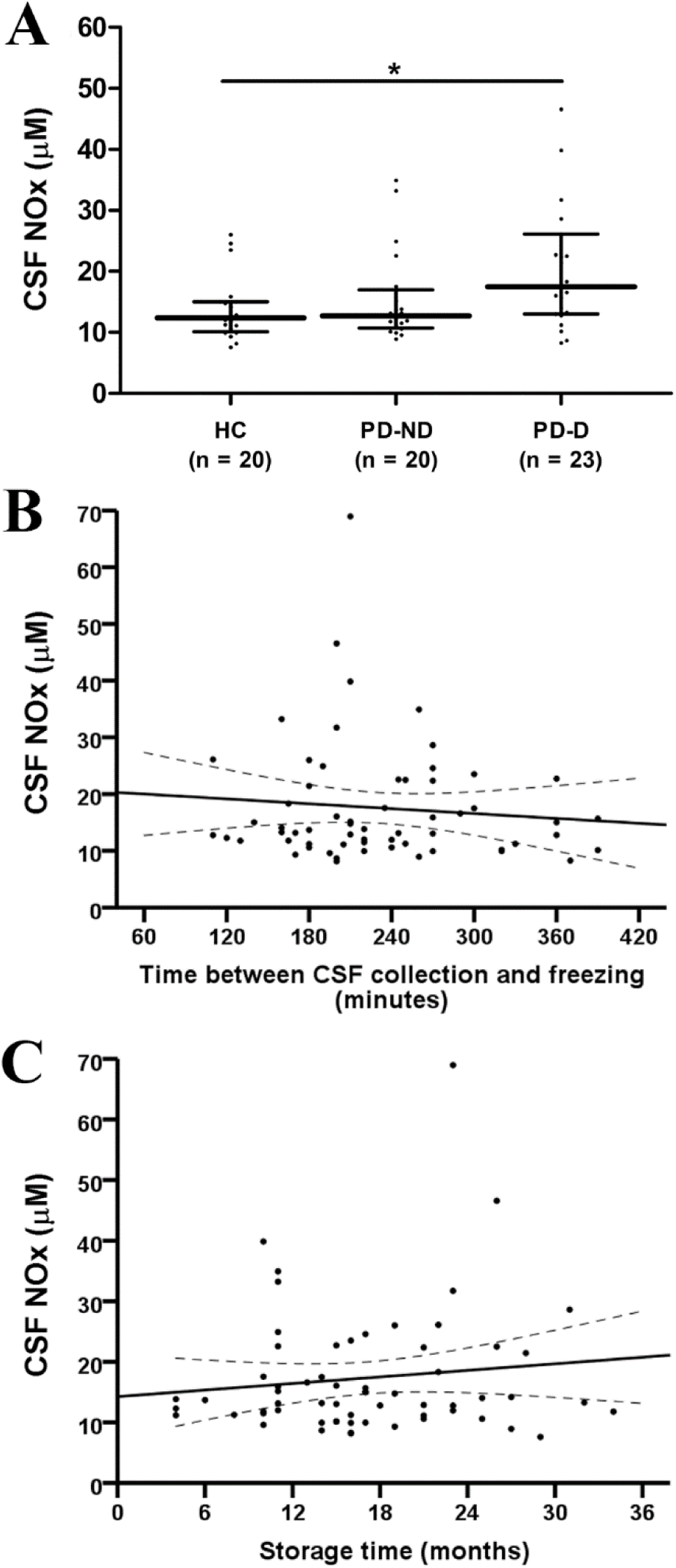
CSF nitrite and nitrate (NOx) levels in patients with Parkinson’s disease (PD) and healthy controls. A: CSF NOx levels between healthy controls, patients with PD without levodopa-induced dyskinesia (LID), and patients with PD with LID. B: Correlation between CSF NOx levels and time between CSF collection and freezing in minutes. C: Correlation between CSF NOx levels and storage in months. * p < 0.05. Brackets represent median and interquartile range. Abbreviations: HC, healthy controls; PD-D, Patients with PD with LID; PD-ND, Patients with PD without LID.

### 3.3. Associations between CSF nitrite and nitrate levels and clinical variables

There was no correlation between CSF NOx levels and age at onset of PD, disease duration, levodopa therapy duration, antiparkinsonian drugs, doses. MDS-UPDRS (total and subitems) and Hoehn and Yahr stage. There was no correlation between CSF NOx levels and items 4.1 (time spent with dyskinesias) and item 4.2 (functional impact of dyskinesias) of MDS-UPDRS Part IV in PD-D group (n = 23). Furthermore, there was no correlation between Part 1B, Part 3 and Part 4, historical subscore, objective subscore and total score of UDysRS.

Median time between sample collection and freezing was 240 minutes (interquartile range 180–305), and median storage time was 21 months (interquartile range 18–25). No participants were excluded due to CSF cell counts and biochemistry. There was no correlation between CSF NOx levels and time between sample collection and freezing (Spearman’s ρ = –0.05, p = 0.67) (Fig. 1B). There was no correlation between CSF NOx levels and storage time (Fig. 1C). There was no correlation between CSF NOx levels and cell counts, proteins and glucose levels in CSF.

## 4. Discussion

Our results showed NOx levels are increased in CSF of patients with PD with LID compared to healthy controls. However, there was no difference between patients with PD with and without LID. CSF NOx levels were not influenced by external factors, as processing and storage time, as well as by CSF cells and proteins.

These results are in agreement with previous authors which reported higher CSF levels of nitrite [7] and summed CSF nitrite and nitrate levels [10] in patients with PD compared to controls. However, other studies showed no differences in CSF nitrite and nitrate levels between patients with PD and controls [8, 9, 11] and even reduced CSF levels of nitrite in PD [6]. In 22 drugnäive patients with PD, Boll, Alcaraz-Zubeldia, Montes and Rios [10] reported a 4-fold increase in CSF NOx compared to controls supposedly caused by oxidative stress.

In the present work, patients with PD and LID presented more severe features than patients without LID. The difference of clinical severity between the two PD groups could affect statistical analyses. However, covariance analysis had showed a more severe disease did not influence CSF NOx levels.

We used an ozone-based chemiluminescence technique to measure NOx, considered as a direct method to detect smaller quantities of NO in real time with high accuracy [14]. None of the previous studies involving NOx measurement in patients with PD employed this technique. Griess reaction, an indirect method with low cost to measure NO by detecting nitrite in samples, was used in many previous studies [6, 7, 8, 9, 11]. However, Griess reaction has some technical issues (external nitrite contamination, sample nitrite degradation, reagent problems) [14], which may explain distinct results between similar works.

As study strengths, we had efficient CSF collection and processing protocols, without external factor influences. Patients with PD and healthy controls were evaluated by the same movement disorders specialist, reducing variability in clinical variables. We used one of the main instruments to evaluate LID (UDysRS) according to the Movement Disorders Society, and our dyskinetic patients had moderate scores in UDysRS. As limitations, we had a small number of participants. The three groups (HC, PD-ND, PD-D) were age-matched, but patients with PD and healthy controls were not sex-matched. The inclusion of a drug-näive group of patients with PD could help to explore CSF NOx levels in untreated patients.

## 5. Conclusions

The present study demonstrated NO is more expressed by the central nervous system in patients with PD and LID. This finding strengthens the role of NO on LID pathophysiology.

## Data Availability

The datasets generated during and/or analyzed during the current study are available from the corresponding author on reasonable request.

## Acknowledgments

We would like to thank Manuelina Macruz Capelari, Nathália Novaretti and Larissa Serveli for technical support.

## Abbreviations

PD – Parkinson’s disease

LID – Levodopa-induced dyskinesias NO – Nitric oxide

CSF – Cerebrospinal fluid

NOx – Nitrite and nitrate

